# The Place of Fluvoxamine in the Treatment of Non-critically ill Patients with COVID-19: A Living Systematic Review and Meta-analysis

**DOI:** 10.1101/2021.12.19.21268044

**Authors:** Salah Eddine Oussama Kacimi, Elona Greca, Mohamed Amine Haireche, Ahmed Sallam ElHawary, Mounir Ould Setti, Rebecca Caruana, Sahar Rizwan, Hidayet Benyettou, Mohammad Yasir Essar, Jaffer Shah, Sherief Ghozy

## Abstract

**Background:** Fluvoxamine is a selective serotonin reuptake inhibitor that is known to be used as antidepressant. Repurposing of Fluvoxamine for the treatment of COVID-19 is theorized to help in the prevention of the clinical deterioration of SARS CoV-2 patients. In our systematic review and meta-analysis, we aim to assess the safety and efficacy of the drug under study in terms of its effect on the mortality and the risk of hospitalization and mechanical ventilation in non-critically ill COVID-19 patients.

**Methods:** We performed a systematic search of seven electronic databases. The search results were screened based on the previously determined inclusion and exclusion criteria. We determined the data related to our objectives. The mortality rates, rates of hospitalization, risk of mechanical ventilation and serious side effects were extracted from the studies that successfully met our inclusion and exclusion criteria. Then, the extracted data from the included studies was included in the meta-analysis.

**Results:** Three studies, two randomized clinical trials and one observational cohort study, with 1762 patients, were the final outcome of our search and screening processes. Among all participants, 886 patients received Fluvoxamine while 876 were controls. Follow up periods ranged from 7 days to 28 days. There was no significant difference in the intention-to-treat mortality rates between the two groups (RR = 0.66; 95% CI: 0.36 - 1.21, p-value = 0.18; *I*^*2*^ = 0%). However, Fluvoxamine decreased the per-protocol mortality compared to both placebo alone or placebo/standard care (RR = 0.09; 95% CI: 0.01 - 0.64, p-value = 0.02; *I*^*2*^ = 0% and RR = 0.09; 95% CI: 0.01 - 0.72, respectively). As compared to placebo or standard care, the all-cause hospitalization was significantly reduced in the fluvoxamine group (RR = 0.71; 95% CI: 0.54 - 0.93, p-value = 0.01; *I*^*2*^ = 61%). This risk reduction was not significant when compared to placebo alone (RR = 0.76; 95% CI: 0.57 - 1.00; p-value = 0.051; *I*^*2*^ = 48%). Furthermore, the risk of mechanical ventilation was not improved in the fluvoxamine group as compared to placebo (RR = 0.71; 95% CI: 0.43 - 1.16, p-value = 0.17; *I*^*2*^ = 0%). The serious adverse effects were almost the same in the treatment group and the control (13% and 12% respectively).

**Conclusion:** Fluvoxamine does not significantly reduce the mortality rates or the risk of mechanical ventilation in SARS CoV-2 patients. Nonetheless, it was found to have a good impact on reducing all cause hospitalization among patients with COVID-19 disease. Therefore, further clinical studies are needed to determine the effectiveness of the drug and its mechanisms of action.

## Introduction

Coronavirus disease 2019 (COVID-19) is a pandemic that has emerged in the city of Wuhan, China, at the end of 2019 and has spread with highly transmissibility all over the globe in 2020^[1],[2],[3]^. Symptoms severity, according to the largest cohort of more than 44,000 persons infected with COVID-19 from China, can range from mild (81%), severe (14%) to critical (5%) ^[4],[5]^. During the critical phase, patients may rapidly deteriorate one week after coronavirus disease onset, which increases the urge to discover drugs that prevents this condition ^[6],[7]^.

In November 2020, a randomized clinical trial, reported by Dr. Lenze and his colleagues assessed the efficacy of Fluvoxamine (FLV) in adult patients with COVID-19 for prevention of clinical deterioration ^[8]^. FLV is a selective serotonin reuptake inhibitor (SSRI), mainly used to treat obsessive-compulsive disorder, but also major depressive disorder and anxiety disorders^[9]^. It has not been FDA-approved for the treatment of any infection since April 2021 when it was added to the US National Institutes of Health (NIH) COVID-19 Guidelines Panel ^[10]^.

Potential mechanisms of action of FLV for COVID-19 are the immune modulation aspect and antiviral effect ^[11,12,13]^. There are four factors that might play a key role in immune modulation. First, the activity of the sigma 1 receptor that could be leading to inositol-requiring enzyme 1alpha - driven (IRE1). Second, the decreased activity of platelet aggregation. Third, decreased activity of mast cells. Fourth, increased level of melatonin by cytochrome P450 inhibition. All of them play a significant role in decreasing inflammation during COVID-19^[11,12,14]^. On the other hand, the anti-viral effect of FLV is focused on the interference with viral entry and endolysosomal viral trafficking ^[11]^.

We aimed to carry out a systematic literature review and meta-analysis of all published RCTs and prospective cohort studies to evaluate the safety and efficacy of Fluvoxamine in non-critically ill patients with confirmed COVID-19 diagnosis, with the goal of helping to clarify the possibility of repurposing of this drug.

## Methods

### Data Source

We followed the PRISMA (Preferred Reporting Items for Systematic reviews and Meta-Analyses) checklist for reporting systematic reviews and meta-analyses ^[15]^. One investigator systematically searched Medline, Google Scholar, Scopus, Opengrey, Clinicaltiral.gov, Cochrane, and MedRxiv from database inception till November 1^st^, 2021 to retrieve studies in humans of the efficacy and safety of Fluvoxamine in the treatment of non-critically ill patients with confirmed COVID-19 diagnosis. The following keywords were used for the retrieval of relevant studies: *“Fluvoxamine”, “COVID”, “COVID-19”, and “Coronavirus”*. Additionally, the reference list of the included studies was scrutinized to retrieve further studies. Finally, we included all original studies with no publication date or language restrictions to not miss any relevant papers.

### Study Selection

Studies were considered relevant if 1) their aim was to assess the efficacy and/or safety of Fluvoxamine in the treatment of non-severe cases of COVID-19 2) they are double-arm studies 3) patients in these studies diagnosed with COVID-19 or its symptomatology is not severe enough to require hospitalization at baseline 4) reported enough data to be used in the meta-analysis. Therefore, we included both observational studies (cohort studies) and clinical trials. Studies conducted among ICU-admitted patients were not considered in this meta-analysis. If a specific dataset has been published more than once, we used the most recent publication. Preprints without peer-reviewed publication were also included. We excluded studies that were not reported as full reports such as conference abstracts and letters unless they provide enough data. Secondary studies including systematic reviews, narrative reviews, and computational studies were excluded from this systematic review. Studies performed on animal models were also excluded.

### Data Extraction & Quality Assessment

The following data were extracted by two independent reviewers from each article: country, study design, number of participants in both Fluvoxamine and control arms, Fluvoxamine dose, and drug-intake duration, duration of the follow-up, percentage of participants lost in follow-up, and description of the reported primary outcomes. Additionally, demographic data of studies’ participants were also extracted. Separately, the outcomes reported in the studies including intention-to-treat mortality, per-protocol mortality, mechanical ventilation, hospitalization, presence of serious adverse events (SAE), and presence of any adverse events (AE) were extracted as raw data (event and total) and not as effect estimates. Grade 3, 4, and 5 adverse events reported by the TOGETHER study ^[16]^ were considered as SAEs. The quality of the included studies was assessed by two other independent reviewers using the NIH Quality Assessment Tool for Controlled Intervention Studies and Cohort Studies ^[17]^.

### Data Synthesis & Analysis

The primary endpoint was to assess the effect of Fluvoxamine on the risk of mortality, mechanical ventilation, or hospitalization among COVID-19 patients without severe illness at baseline. In the first part, we gathered raw data represented by the number of event occurrences among total patients in each arm from both observational and interventional studies. Thereafter, we calculated pooled effect estimates and their corresponding 95% confidence intervals (CI) from these raw data. At the second time, only interventional studies comparing Fluvoxamine with placebo were considered in separate meta-analyses. Because of the prospective nature of all the included studies, Relative Risk (RR) was used as an effect estimate.

All data were analyzed by means of the “meta” package; R software version 4.0.2. We computed the pooled RRs and their corresponding 95% CI using the fixed effects model. Heterogeneity of the effect across studies was assessed by Q^2^ statistics, which is distributed as *X*^2^ statistics ^[18]^. A value of p-value < 0.05 was used to indicate a lack of homogeneity (heterogeneity) among effects. *I*^*2*^ statistics were provided to quantify the percentage of total variation across studies that was attributable to heterogeneity rather than to chance. We used a fixed-effects model if the *I*^*2*^ p-value significance was < 0.05. Otherwise, a random-effects model was used. Due to the small number of the included studies (<10 per the analysis), neither subgroup analysis, Egger’s regression test for the assessment of publication bias nor meta-regression were possible ^[19,20]^.

## Results

### Inclusion flowchart

Two independent reviewers screened the title and abstracts of 155 potentially relevant non-duplicate studies. Of the 155 abstracts, a total of 96 abstracts was excluded due to the reasons mentioned in the PRISMA flow diagram (**Figure 01**). Consequently, full texts of 11 studies were assessed for eligibility of which 8 were excluded (7 studies did not meet the first eligibility criteria and 1 study was reported among patients admitted to the ICU ^[21]^. Additionally, no studies were identified through hand-searching of the reference list of the included studies. Finally, three studies (2 interventional and 1 observational) met the eligibility criteria and were included in the review and meta-analysis. The PRISMA flow diagram is depicted in (**Figure 01**)

**Figure 01:**
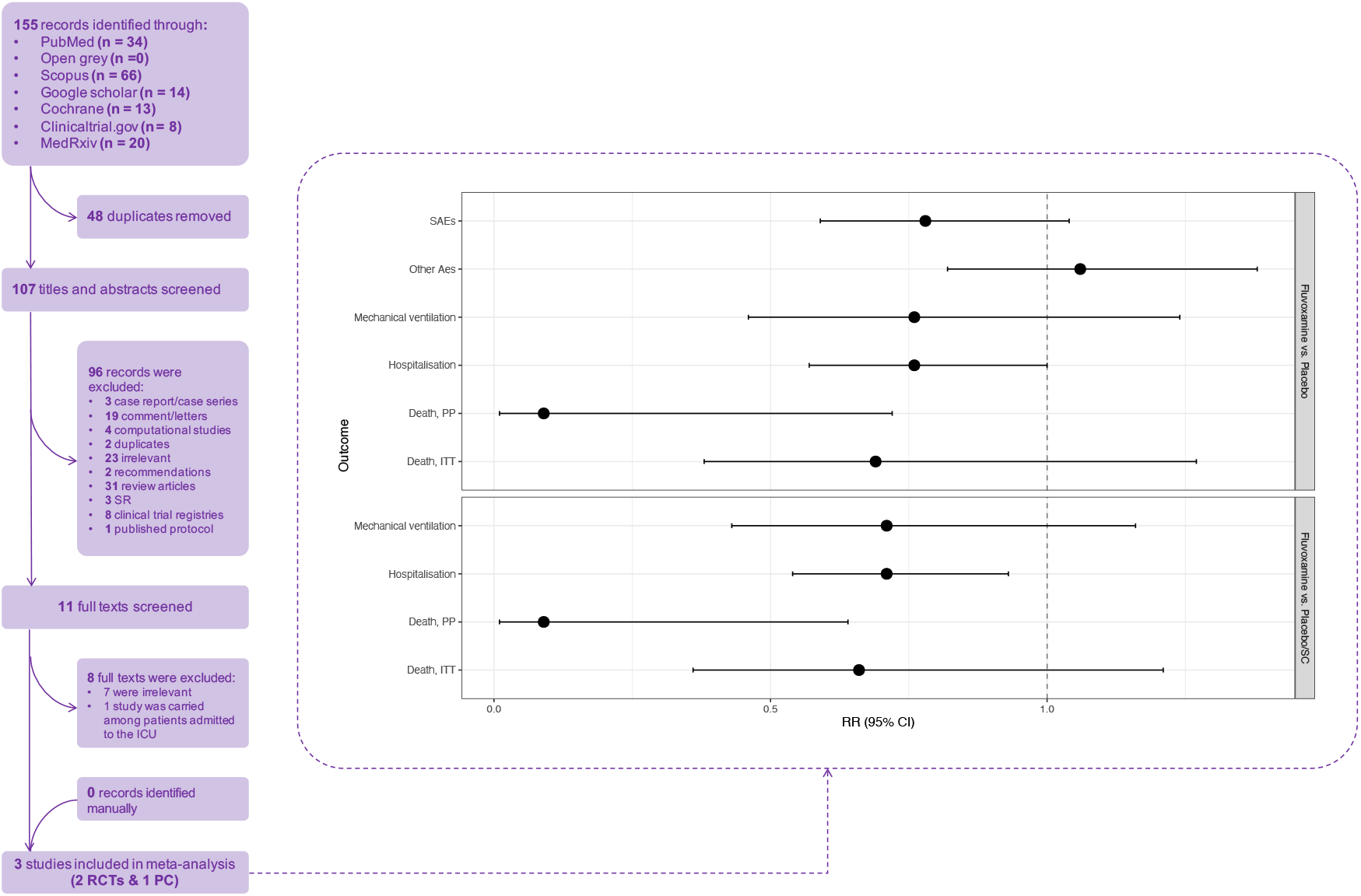
PRISMA flow chart of the systematic review.

### Description of the included studies

All included studies were published in English. Among the three studies, two ^[16,8]^ were randomized double-blinded placebo-controlled trials and one ^[22]^ was an observational cohort study. All the included studies were prospective with a maximum follow-up duration of 28 days reported in the TOGETHER study. The minimum follow-up duration was 14 days. The sample size of the included studies ranged from 133 to 1497. Two studies were conducted in the United State of America and one study in Brazil. Fluvoxamine was administered daily at different doses and durations in the included studies. Primary outcomes were heterogeneous among included studies. COVID OUT study assessed the clinical deterioration of COVID-19 patients, TOGETHER study assessed the retention in COVID-19 emergency or transfer to tertiary care, while Seftel et al. assessed mortality, ICU admission, and persistence of COVID-19 symptomatology as the primary outcome. Details of the characteristics of the included studies can be found in (**Table1**)

**Table 01:**
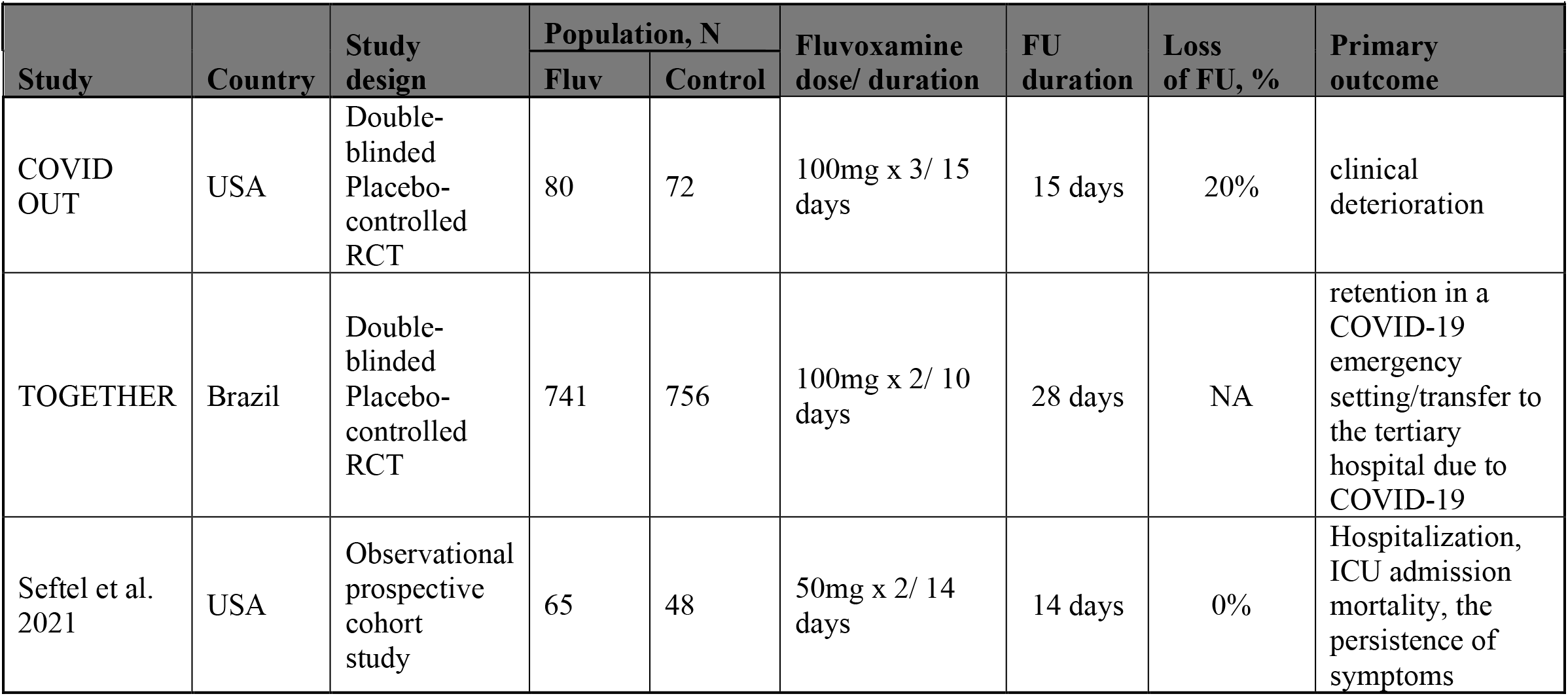
Characteristics of the included studies.

| Table 01: Characteristics of the included studies |  |  |  |  |  |  |  |  |
| --- | --- | --- | --- | --- | --- | --- | --- | --- |
| Study | Country | Study design | Population, N |  | Fluvoxamine dose/ duration | FU duration | Loss of FU, % | Primary outcome |
|  |  |  | Fluv | Control |  |  |  |  |
| COVID OUT | USA | Double-blinded Placebo-controlled RCT | 80 | 72 | 100mg x 3/ 15 days | 15 days | 20% | clinical deterioration |
| TOGETHER | Brazil | Double-blinded Placebo-controlled RCT | 741 | 756 | 100mg x 2/ 10 days | 28 days | NA | retention in a COVID-19 emergency setting/transfer to the tertiary hospital due to COVID-19 |
| Seftel et al. 2021 | USA | Observational prospective cohort study | 65 | 48 | 50mg x 2/ 14 days | 14 days | 0% | Hospitalization, ICU admission mortality, the persistence of symptoms |

### Baseline characteristics of studies’ participants

All the included studies reported age, percentage of gender, ethnicity/race, and comorbidities. However, Seftel et al. failed to provide data regarding body-mass index (BMI). Additionally, Seftel et al. reported data on only three comorbidities (hypertension, diabetes, and chronic lung disease). Baseline characteristics were not different between Fluvoxamine arm and control arm among the included studies except for gender that was not well-matched in Seftel et al. with 59% of males in Fluvoxamine arm and 41% in standard care arm. Diabetes, hypertension, and lung disease are the most prevalent comorbidities among participants. Full demographic data can be found in (**Table2**)

**Table 2:** Baseline characteristics among patients of the included studies.

|  |  |  |  |  |  |  |
| --- | --- | --- | --- | --- | --- | --- |
| Baseline Characteristics | TOGETHER |  | COVID OUT |  | Seftel et al. 2021 |  |
|  | Fluvoxamine<br>(N = 741) | Placebo<br>(N = 756) | Fluvoxamine<br>(n = 80) | Placebo<br>(n = 72) | Fluvoxamine<br>(n = 65) | Standard care<br>(n = 48) |
| Age, Median (IQR) | 50 (39–56) | 49 (38–56) | 46 (35-58) | 45 (36-54) | 44 ± 15<sup>a</sup> | 43 ± 15<sup>a</sup> |
| Males, % | 45 | 40 | 30 | 26 | 59 | 41 |
| Race, % |  |  |  |  |  |  |
| Black or African American | 1 | 1 | 23 | 28 | 1.5 | 0 |
| Mixed race | 96 | 95 | - | - | - | - |
| White | 1 | 1 | 70 | 69 | 5 | 27 |
| Unknown | 3 | 3 | 1 | 0 | - | - |
| Ethnicity, % |  |  |  |  |  |  |
| Asian | - | - | 4 | 1 | 0 | 2 |
| Latino | - | - | 4 | 3 | 94 | 71 |
| Chronic comorbidity, % |  |  |  |  |  |  |
| Anxiety | - | - | 6 | 1 | - | - |
| Asthma | 2 | 2 | 21 | 13 | - | - |
| Auto-immune diseases | 0 | < 1 | - | - | - | - |
| Chronic heart disease | 1 | 1 | - | - | - | - |
| Chronic kidney disease | < 1 | < 1 | - | - | - | - |
| Chronic neurological disease | 1 | 1 | - | - | - | - |
| Chronic pulmonary disease | 1 | < 1 | - | - | 3 | 2 |
| Depression | - | - | 1 | 6 | - | - |
| Diabetes | 17 | 15 | 11 | 11 | 17 | 8 |
| Hyperlipidemia | - | - | 9 | 10 | - | - |
| Hypertension | 14 | 12 | 19 | 21 | 17 | 35 |
| Rheumatological disorder | < 1 | 0 | 5 | 4 | - | - |
| Any other disease | 3 | 3 | - | - | - | - |
| Body-mass index, % |  |  |  |  |  |  |
| <30 kg/m2 | 48 | 49 | 46 | 42 | - | - |
| ≥30 kg/m2 | 51 | 50 | 54 | 58 | - | - |
| Unspecified | 1 | 1 | 0 | 0 | - | - |
| Abbreviations: IQR, interquartile range; a, mean ± SD. All results are displayed as percentages. |  |  |  |  |  |  |

### Outcomes

#### Fluvoxamine compared to placebo or standard care (observation)

Fluvoxamine did not reduce risk of mortality from COVID-19 RR = 0.66 (95% CI: 0.36 - 1.21, p-value = 0.18; *I*^*2*^ = 0%). However, when we used the per protocol mortality data of TOGETHER study, Fluvoxamine has significant reduction in per protocol mortality as compared to the placebo or standard care (SC) RR = 0.09 (95% CI: 0.01 - 0.64, p-value = 0.02; *I*^*2*^ = 0%). As compared to placebo or SC, Fluvoxamine showed benefit on reducing risk for all-cause hospitalization RR = 0.71 (95% CI: 0.54 - 0.93, p-value = 0.01; *I*^*2*^ = 61%), but did not for the risk of mechanical ventilation (MV) RR = 0.71 (95% CI: 0.43 - 1.16, p-value = 0.17; *I*^*2*^ = 0%). Forest plots can be found on the (**Supplementary**).

#### Fluvoxamine compared to a placebo alone

When compared to placebo alone, Fluvoxamine did no reduce the risk of intention-to-treat death RR = 0.69 (95% CI: 0.38 - 1.27, p-value = 0.24), but showed a benefit on per protocol death RR = 0.09 (95% CI: 0.01 - 0.72, p-value = 0.02). When compared to placebo alone, Fluvoxamine did not reduce the risk for both MV and hospitalization RR = 0.76 (0.46 - 1.24, p-value = 0.27; *I*^*2*^ = 0%) and RR = 0.76 (0.57 - 1.00, p-value = 0.051; *I*^*2*^ = 48%), respectively. Incidence of serious adverse events was not different among Fluvoxamine (10%) and placebo arm (12%) (p-value for RR = 0.09). Similarly, the incidence of any other adverse events was almost identical between treatment group (13%) and placebo group (12%) RR = 1.06 (0.82 - 1.38, p-value = 0.65; *I*^*2*^ = 0%). Details regarding outcomes can be found on (**Table3**)

**Table 3:**
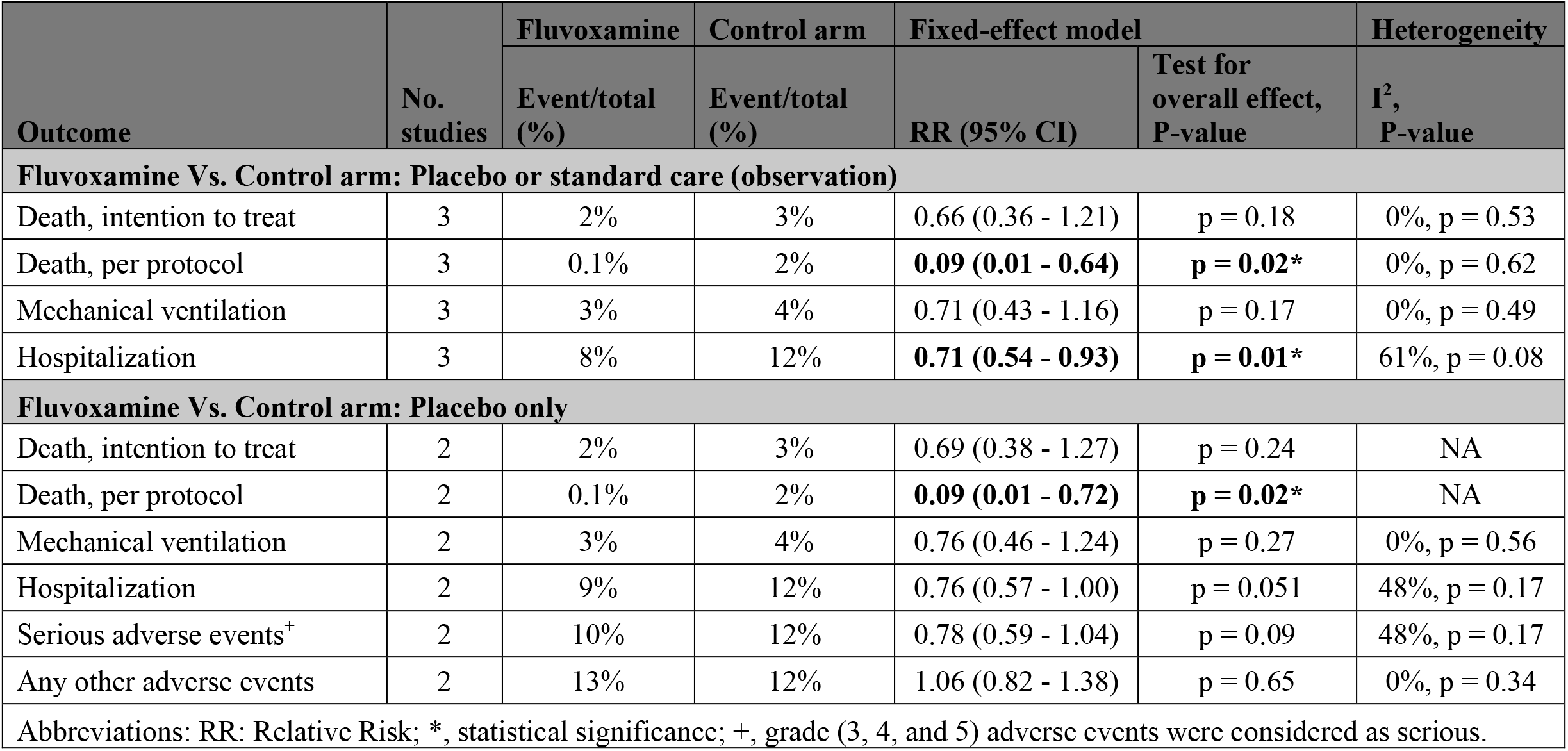
Summary of the patients’ outcomes.

### Risk of bias

Using the NIH quality assessment tool for interventional studies, we find both RCTs to be of good quality. On the other hand, the quality of the prospective cohort study by Seftal et al. was fair using the same tool adapted for cohort studies.

## Discussion

COVID-19 disease is the disease of the era. Its high prevalence forced the Scientists to race against time to find a vaccine that prevents infection, as well as a treatment for the severe symptoms of the disease. Many theoretical experiments have suggested the repurposing of particular drugs, and accordingly, many clinical studies have been conducted to verify the effectiveness of these proposed drugs ^[23,24]^. Some of these medicines have proven effective, whether in relieving the symptoms or even in curing the disease. Fluvoxamine was one of these drugs ^[10]^. It is a drug for the nervous system used as an antidepressant through its action as a selective serotonin reuptake inhibitor (SSRI) ^[25]^. The use of antidepressant drugs as antiviral agents was not of the moment. They have previously been used in the Middle East Respiratory Syndrome (MERS) and the Severe Acute Respiratory Syndrome (SARS), and some clinical and preclinical studies have proven their success in improving the condition of patients with these two diseases ^[26,27]^.

Repurposing of fluvoxamine was theorized to be beneficial in preventing the progression of severity of SARS CoV-2 disease ^[8]^. These theories were based on the anti-inflammatory properties of the drug which come mainly from its moderate to high affinity to sigma-1 receptors (S1R) ^[28]^. Among all antidepressants, fluvoxamine was found to have the highest potency at S1R ^[29-31]^. S1R is an endoplasmic reticulum (ER) chaperone protein that plays a regulatory role of cytokines production in the sittings of inflammation ^[30]^. It interferes with the ER stress (cytokine storm) syndrome which occurs accordingly to the SARS CoV-2 viral replication within the human cells and contributes to the high mortality of the disease ^[30,32-33]^. In addition, S1R was identified by Gordon et al to play a pivotal inhibitory role in the viral RNA replication process of SARS CoV-2 ^[30,34]^. Thus, the agonistic effect of fluvoxamine on sigma-1 receptors has supported the theories of its use as anti-inflammatory and antiviral agent in the treatment of SARS CoV-2 disease ^[29]^. There are many other theories (e.g. inhibition of platelet aggregation, mast cell degranulation and lysosomotropism) linking the mechanisms of action of fluvoxamine and its possible efficacy in COVID-19 patients ^[11]^.

A few clinical studies have been conducted to confirm the efficacy of fluvoxamine in alleviating the severe symptoms and preventing the progression of the disease. In our systematic review and meta-analysis, we assessed the efficacy of fluvoxamine in terms of its effects on mortality rates, risk of hospitalization and mechanical ventilation as factors that can be relied upon to determine the effectiveness of the drug in treating the disease.

Although the COVID OUT randomized clinical trial (which was performed by Lenze et al ^[8]^) was not a large trial, it opened the gaze for the high need to perform larger trials to reach a reliable conclusion about the efficacy of the drug. Of all the included studies, the TOGETHER trial was the largest and the most recent trial ^[16]^.

The increased mortality rates among COVID 19 patients increased the demand to check for drugs that may reduce the high risk of death especially in advanced age ^[35, 36]^. In their large multicenter retrospective observational cohort study, Oskotsky et al indicated the effectiveness of SSRIs, specifically fluoxetine and fluoxetine or fluvoxamine, in reducing the mortality rates among the groups that used this category of antidepressants compared to the control group ^[37]^. In our review, we found that there was no significant difference in the ITT mortality rates between fluvoxamine group and placebo or SC (RR = 0.66; 95% CI: 0.36 - 1.21). However, the PP mortality from the TOGETHER trial indicated a real and effective difference between the two groups (RR = 0.09; 95% CI: 0.01 - 0.64). Therefore, this can give the preference to the use of fluoxetine over fluvoxamine in the reduction of mortality among COVID-19 patients. We can also infer from this the superiority of fluoxetine and fluvoxamine over the other SSRIs ^[37, 38]^.

## Conclusion

Repurposing of fluvoxamine in treatment of SARS CoV-2 patients has not shown significant efficacy in reducing either the mortality rates or mechanical ventilation. However, it showed promising results in reducing the risk for hospitalization among patients with COVID-19 disease. There is still an urgent need for more clinical studies to determine the extent of its effectiveness and to know more about its biological mechanisms in COVID-19 patients.

## Data Availability

All data produced in the present study are available upon reasonable request to the authors
All data produced in the present work are contained in the manuscript
All data produced are available online at PubMed

